# Integrating a Polygenic Risk Score for Coronary Artery Disease as a Risk Enhancing Factor in the Pooled Cohort Equation is Cost-effective in a US Health System

**DOI:** 10.1101/2021.06.21.21259210

**Authors:** Deo Mujwara, Geoffrey Henno, Stephen T Vernon, Siyang Peng, Paolo Di Domenico, Brock Schroeder, George B Busby, Gemma A Figtree, Giordano Bottà

**Affiliations:** Allelica, Inc; Illumina, Inc; Kolling Institute, Royal North Shore Hospital, Sydney, NSW, Australia; Charles Perkins Centre, University of Sydney, Sydney, NSW, Australia; Department of Cardiology, Royal North Shore Hospital, Sydney, NSW, Australia

## Abstract

**Importance:** The pooled cohort equation (PCE) is used to determine an individual’s 10-year risk (low, borderline, intermediate, or high) of atherosclerotic cardiovascular disease (ASCVD) but it fails to identify all individuals at high risk. Those with borderline or intermediate risk require additional risk enhancing factors to guide preventive therapy decisions. Including a polygenic risk score (PRS) for coronary artery disease as a risk enhancing factor improves precision in determining the risk of ASCVD and informs decisions for prevention therapy.

**Objective:** To assess the cost-effectiveness of integrating PRS for coronary artery disease with the PCE to determine an individual’s 10-year risk for ASCVD compared to the PCE-alone.

**Design, setting, and population:** A Markov model was developed on a hypothetical cohort of 40-year-old individuals in the US with borderline or intermediate PCE 10-year risk for ASCVD who fall in the top quintile of the PRS distribution and are not on preventive therapy (e.g., statins). Model transition probabilities and economic costs came from existing literature with costs reflecting a payer perspective and inflation-adjusted to 2019 US$.

**Interventions:** The modeled strategies were: (1) the PCE-alone and (2) the PCE with PRS for coronary artery disease as a risk enhancing factor. Analyses were performed at 5 year, 10 year, and lifetime time horizons.

**Main outcomes and measures:** Quality-adjusted life-years (QALYs) gained, acute coronary syndromes and ischemic stroke events prevented, mean costs, and incremental cost-effectiveness ratios (ICER) were measured. One-way, two-way, and probabilistic sensitivity analyses were used to assess uncertainty in parameter estimates. Future costs and health benefits were discounted at an annual rate of 3%.

**Results:** Compared to the PCE-alone, PCE+PRS was cost-saving, effective and cost-effective (dominant). A health system would save more than $500, $2,300, and $9,000 per additional high-risk individual identified using PCE+PRS and prevent 27, 47 and 83 acute CAD or ischemic stroke events per 1,000 persons in 5 year, 10 year, and lifetime time horizons, respectively.

**Conclusions and relevance:** Implementing PRS as a risk enhancing factor for CAD among individuals with borderline or intermediate 10-year risk reclassifies individuals as high-risk who would otherwise remain unidentified, prevents future acute CAD and ischemic stroke events, and both saves money and is cost-effective for health systems.

**Key Points:** *Question:* Is it cost-effective to use polygenic risk scores (PRS) for coronary artery disease (CAD) among individuals with borderline or intermediate risk of atherosclerotic cardiovascular disease (ASCVD) to inform preventive therapy decisions?

*Findings:* We modeled a hypothetical cohort of individuals with borderline or intermediate risk of ASCVD who fall in the top quintile of the CAD-PRS distribution but not on preventive therapy. Integrating CAD-PRS in the pooled cohort equation improved quality-adjusted life-years, saved money and was cost-effective.

*Meaning:* Integrating PRS as an enhancing factor in the pooled cohort equation risk assessment for ASCVD used in current clinical practice was cost-effective.

## Introduction

The pooled cohort equation (PCE) is used to determine an individual’s 10-year risk of atherosclerotic cardiovascular disease (ASCVD) but it does not identify all individuals at high risk^1,2^ leading to missed opportunies to intervene and prevent adverse health outcomes. Strong evidence shows a substantial proportion of coronary artery disease (CAD) is attributable to genetic factors^3–6^ which are not currently considered in the PCE. The integration of such genetic risk factors into the primary prevention setting remains limited and the cost-effectiveness is unknown.

To guide preventive therapy interventions the PCE 10-year risk for ASCVD stratifies individuals into four risk categories: low (<5%), borderline (5% to <7.5%), intermediate (≥7.5% to <20%), and high (≥20%).^7^ Statin therapy is effective in preventing CAD^8^ and recommended for high-risk individuals.^7^ But, for those with borderline or intermediate risk, the presence of additional risk-enhancing factors, which by definition increase ASCVD risk by at least two fold, is needed to guide preventive therapy decisions.^7^ Previous work has shown increased 30-day all-cause mortality and worse health outcomes in patients with ST-elevation myocardial infarction (STEMI) in the absence of standard clinical cardiovascular risk factors used in the PCE (e.g., hypercholesterolemia, diabetes, and smoking) compared to those with risk factors,^9^ indicating an urgent need to improve the risk models used to determine ASCVD risk and to guide preventive therapy.

Polygenic risk scores (PRS) for CAD have been shown to be strong independent predictors of disease.^1,4,10^ PRS are developed using large populations and clinical biobanks and integrate the number of risk variant alleles for an individual weighted by the impact of each allele on disease risk.^11,12^ Individuals who fall in the top quintile of the CAD-PRS distribution have around a 2-fold increased risk of CAD events compared to the remainder of the population (odds ratio [95% confidence interval]: 1.9[1.8-2.0]^1^ and 2.5[2.4-2.6]^10^). As such, PRS have been proposed as an additional risk enhancing factor for the PCE to improve precision in determining an individual’s 10-year risk, particularly among those with borderline or intermediate risk.^1^

Establishing the cost-effectiveness of CAD-PRS in a clinical setting may encourage payers to support this genetic testing in standard clinical practice. Previous studies have found using genetic testing as an additional risk factor in screening for chronic conditions (e.g. breast cancer) cost-effective,^13,14^ including use of PRS,^15–17^ but little is known on how PRS can inform preventive therapy for CAD in clinical practice.^18,19^ Here, we examined the cost-effectiveness of implementing CAD-PRS among individuals with borderline or intermediate PCE 10-year risk to reclassify those in the top quintile of the PRS distribution as high CAD risk and who are thus eligible for, but currently not prescribed, preventive therapy.

## Methods

### Overview and Model Structure

We developed a Markov model to project costs and quality-adjusted life years (QALYs) in a cohort of individuals ≥ 40 years at high-risk of CAD. The model had an annual cycle length with health states (**Figure 1**) defined to reflect occurrence of acute CAD (acute myocardial infarction), acute ischemic stroke, statin adherence, statin side effects and mortality (not shown in Figure 1). Parameter inputs were taken from published sources with costs estimated from a payer perspective and inflation adjusted to 2019 US$. The relative performance of strategies was assessed using the incremental cost-effectiveness ratio (ICER), expressed in US$/QALY gained, and the cost-effectiveness determined according to the willingness to pay (WTP) threshold equivalent to $50,000.^20^ Future costs and QALYs were discounted at an annual rate of 3%. Uncertainty in parameter inputs was assessed using one-way, two-way, and probabilistic sensitivity analyses. Analyses were performed at 5 year, 10 year, and lifetime time horizons using TreeAge Pro Software 2021.

**Figure 1:**
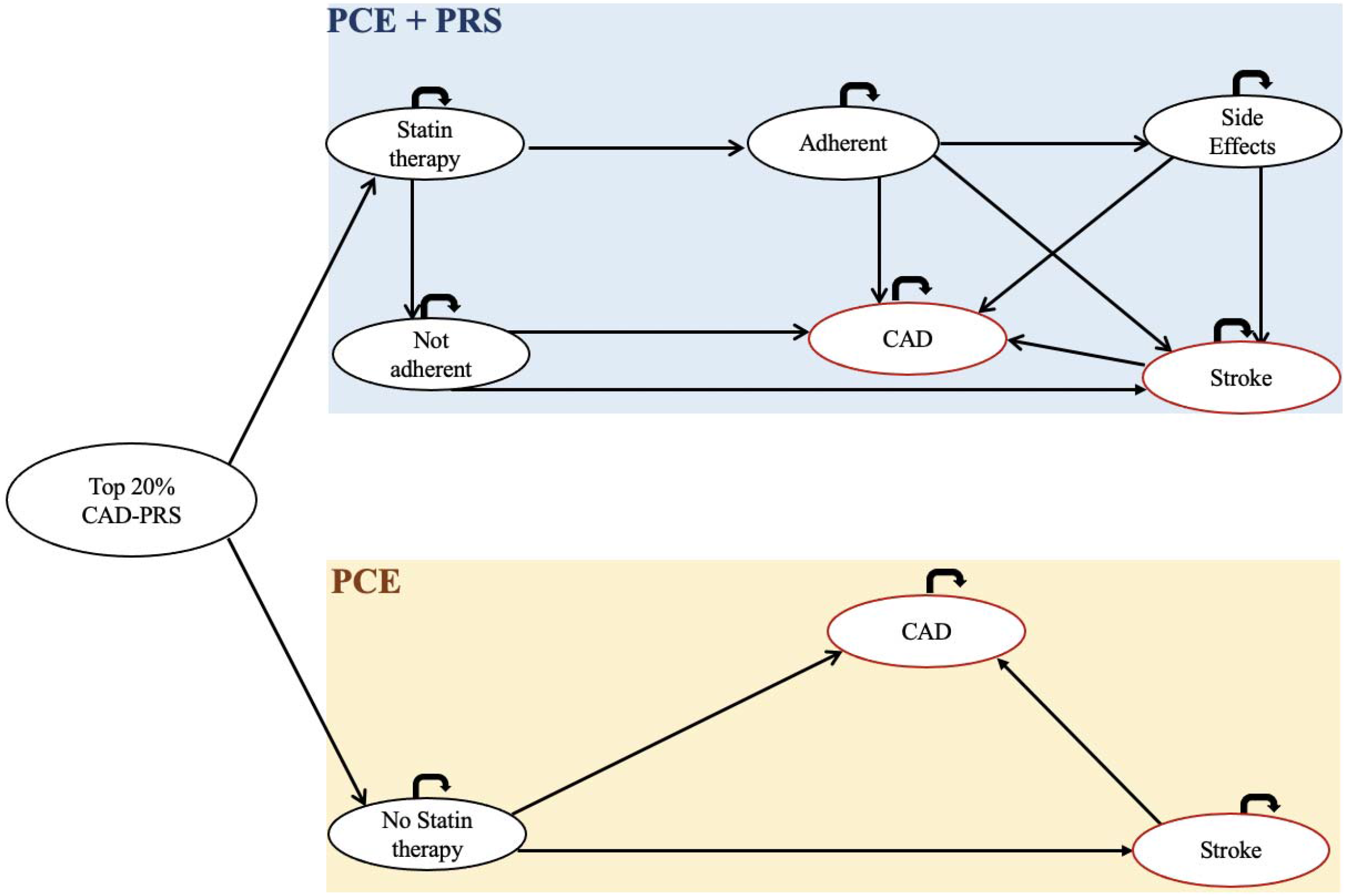
Model Structure*. Abbreviations: PCE = Pooled cohort equation; PRS = Polygenic risk score; CAD = coronary artery disease *The circles represent health states and those indicated with red are the health states for key outcome events (CAD and ischemic stroke). The arrows represent transitions of the cohort across health states or remain the same health state per model cycle.

### Study Population

We applied CAD-PRS on a cohort with borderline or intermediate PCE 10-year risk of ASCVD. Based on Aragam et al. (2020),^1^ 11.1% of individuals with borderline or intermediate 10-year risk will fall in the top quintile of the CAD-PRS distribution. These individuals are not on any preventive therapy and are therefore at high risk of CAD but remain invisible to current clinical ASCVD risk assessment. In this study, the initial cohort represented these high-risk individuals who are currently unidentified by the PCE-alone.

### Strategies

We compared two strategies: i) PCE-alone and ii) PCE+PRS. The PCE-alone strategy represented the current clinical practice that uses conventional risk factors (sex, race, age, blood pressure, lipids, diabetes and smoking status) to determine an individual’s 10-year risk for a first ASCVD,^7^ while the PCE+PRS strategy includes the same risk factors as the PCE-alone strategy with the addition of the CAD-PRS.^1^ Preventive therapy—simvastatin 20-80mg, the most used statin in the US (42% of all statin prescriptions)—was initiated in the PCE+PRS strategy to prevent acute CAD and ischemic stroke events.^21^ In the PCE-alone strategy, we assumed individuals remained unidentified over the analytic time horizon without initiating any preventive therapy. We acknowledge that over the analytical time horizon, age may inform preventive therapy decisions in the PCE-alone strategy^22^ although the impact of age on CAD is likely limited in short time horizons (e.g., 5 to 10 years).

### Model Parameters

Parameter inputs used in the model are listed in **Table 1**. We used a conservative (20%) 10-year risk of ASCVD for individuals classified as high-risk by the PCE-alone^7^ to derive an annual probability of an acute CAD event. Statin therapy effectiveness in reducing the risk of acute CAD came from a randomized controlled trial (RCT) of individuals in the top quintile of the CAD-PRS distribution.^23^ Due to data limitations, the risk of acute ischemic stroke was assumed to be equivalent to that in the general population with a reduced risk among those on statin therapy.^24^ Since no studies have examined the efficacy of simvastatin among individuals with high CAD-PRS, we instead used the efficacy of pravastatin among individuals in the top quintile of the CAD-PRS distribution.^24^ Simvastatin has been shown to be better or comparable to pravastatin in reducing low-density lipoprotein cholesterol.^25,26^ Although adherence to statin in primary prevention tends to be low (<50%) and decreases overtime,^27^ we assumed 50% of the cohort consistently used statins given evidence of higher (80%) adherence among US adults.^28^ Statin adverse effects included myopathy, diabetes and hemorrhagic stroke.^29^ The probability of death in high-risk individuals without CAD, stroke or statin side-effects was derived from social security life tables.^30^ QALY utility weights from the literature were assigned to health states indicating CAD,^31^ stroke,^31^ and statin side effects.^31,32^ Utility weights for high-risk individuals without CAD, stroke, or statin side effects accounted for utility decrement associated with age.^33^

**Table 1:**
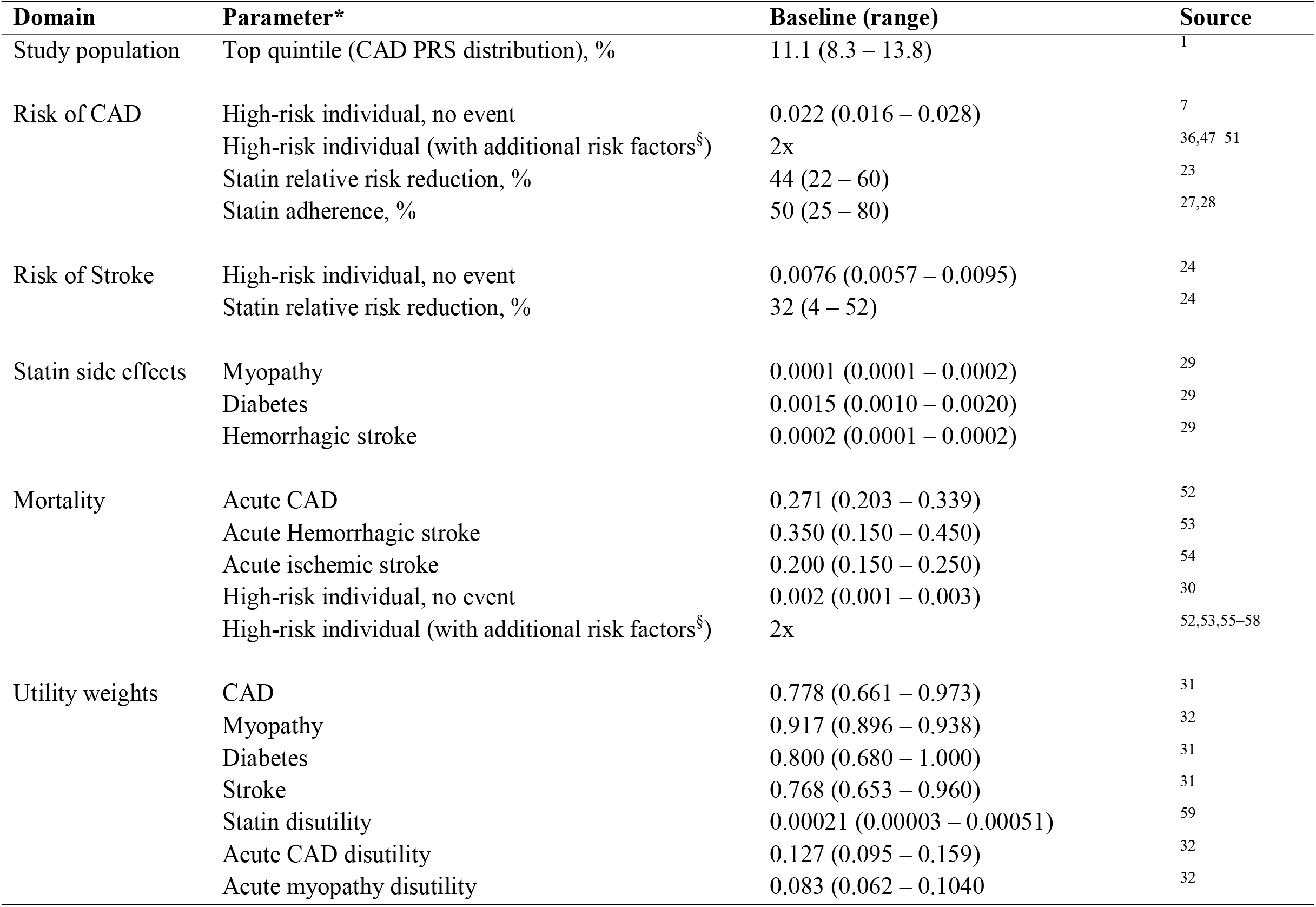
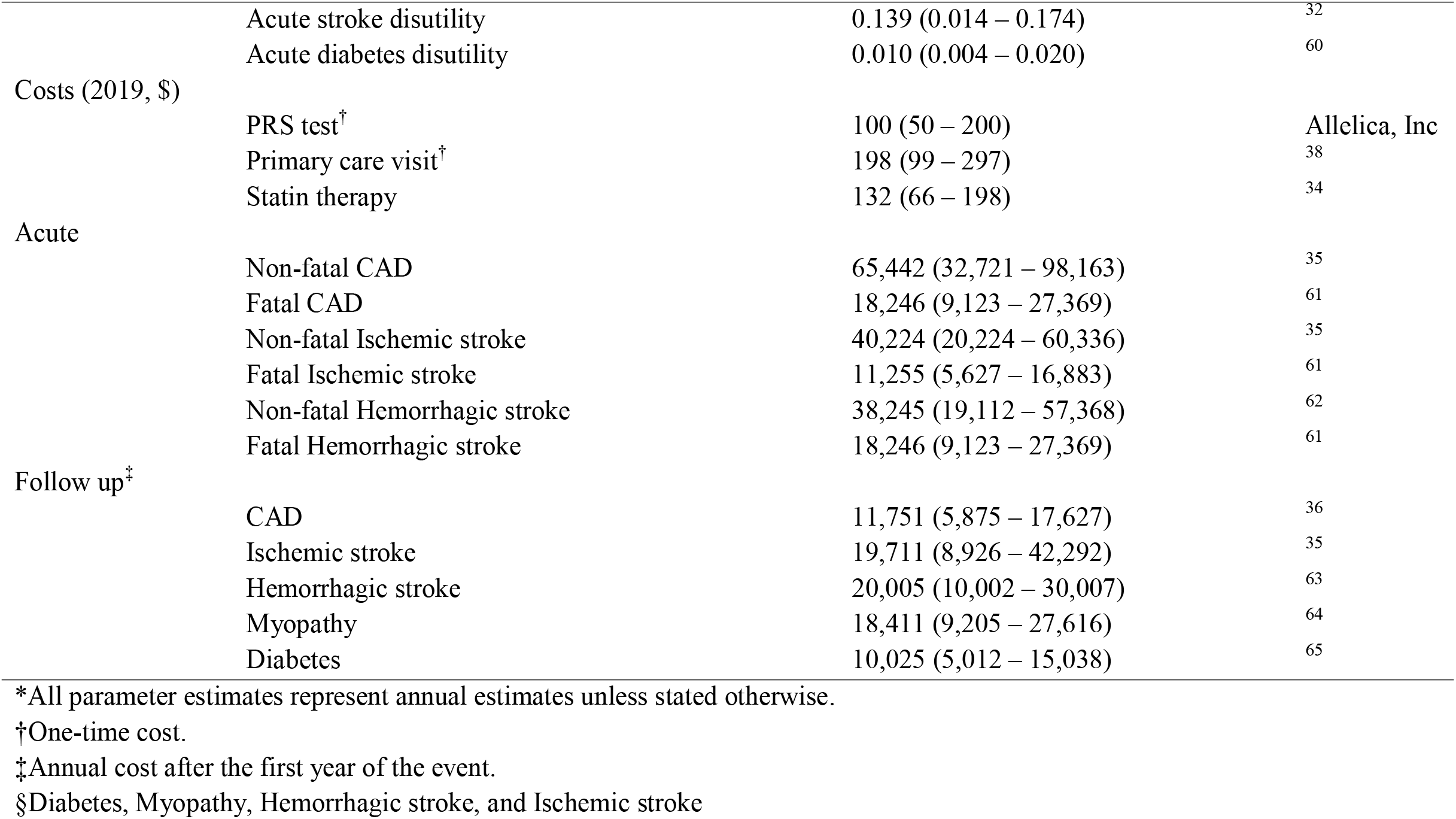
Model parameters.

### Costs

We considered only direct medical costs (Table 1). Costs included: genetic testing, statin therapy, treatment for statin-induced side effects, in-patient hospitalization for fatal and non-fatal acute CAD and ischemic stroke events, and follow-up costs after hospital discharge. The one-time cost ($100) for genetic testing at scale came from Allelica Inc. The cost of statin therapy was derived from online pharmacy prices, which is consistent with the literature.^34^ Costs incurred for treating acute CAD and ischemic stroke were derived from a systematic review of direct medical costs associated with major cardiovascular conditions including in the US.^35^ Costs incurred in treating fatal and non-fatal acute CAD or stroke (ischemic and hemorrhagic) and treatment during the year after hospital discharge were higher compared to subsequent years. Annual costs post an acute CAD event came from claims for commercially insured US adults.^36^

### Base case analysis

We estimated costs and QALYs per strategy with differences in costs (incremental cost, US$) and QALYs (QALY gained) between strategies used to calculate the ICER (US$/QALY gained). The WTP threshold of $50,000 was compared with the ICER for PCE+PRS, such that if the ICER < WTP threshold, PCE+PRS was regarded as cost-effective. We estimated acute CAD and ischemic stroke events, and cost-savings per event prevented by PRS+PRS vs PCE-alone.

### Sensitivity analysis

One-way, two-way, and probabilistic sensitivity analyses were used to assess uncertainty in parameter inputs. One-way sensitivity identified the main cost drivers of variation in the ICER across a range of parameter estimates including costs: PRS testing, statin therapy and treatment for CAD, stroke, and statin side-effects; and probabilities: statin adherence, statin effectiveness, death and getting CAD, ischemic stroke, or statin-effects. The cost of PRS testing and adherence to statin were key drivers of variation in the ICER in the 5-year time horizon, so in the two-way sensitivity, we assessed the impact of the cost of PRS testing on the cost-effectiveness of PCE+PRS at different thresholds (25%, 50%, 75% and 100%) of statin adherence. For probabilistic sensitivity, we assumed beta and gamma distributions for probability and cost parameter inputs, respectively.^37^ We performed 10,000 Monte Carlo simulations and results were reported using cost-effectiveness planes and cost-effectiveness acceptability curves.

We examined various scenarios: first, we assessed the impact of costs involved in getting individuals to undergo genetic testing. We assumed an additional primary care visit to explain to the patient the benefits for PRS and address their concerns.^38^ This cost was not considered in the base case analysis since patients can concurrently get information regarding the benefits of PRS during their initial screening under the PCE-alone strategy, hence the additional primary care visit may not be required. Second, we assumed a 5% annual relative increase in the risk of CAD to account for the impact of age on the risk of cardiovascular diseases. Third, we modeled a best-case scenario with perfect statin adherence and age-adjusted risk for CAD. Finally, we examined the cost-effectiveness of implementing PRS at different start ages (40-75 years) of the cohort.

## Results

### Base case analysis

In the base case (**Table 2**), PCE+PRS compared to PCE-alone was effective (0.02, 0.05 and 0.35 QALYs gained per person), cost-saving ($528, $2,261, and $9,083 per person) and cost-effective (dominant) in 5 year, 10 year, and lifetime time horizons, respectively. Compared to PCE-alone, the PCE+PRS strategy prevented approximately 27, 47 and 83 acute CAD or ischemic stroke events per 1,000 persons, with an average of $19,555, $48,106, and $109,433 saved per event prevented in 5 year, 10 year, and lifetime time horizons, respectively (**Table 3**).

**Table 2.**
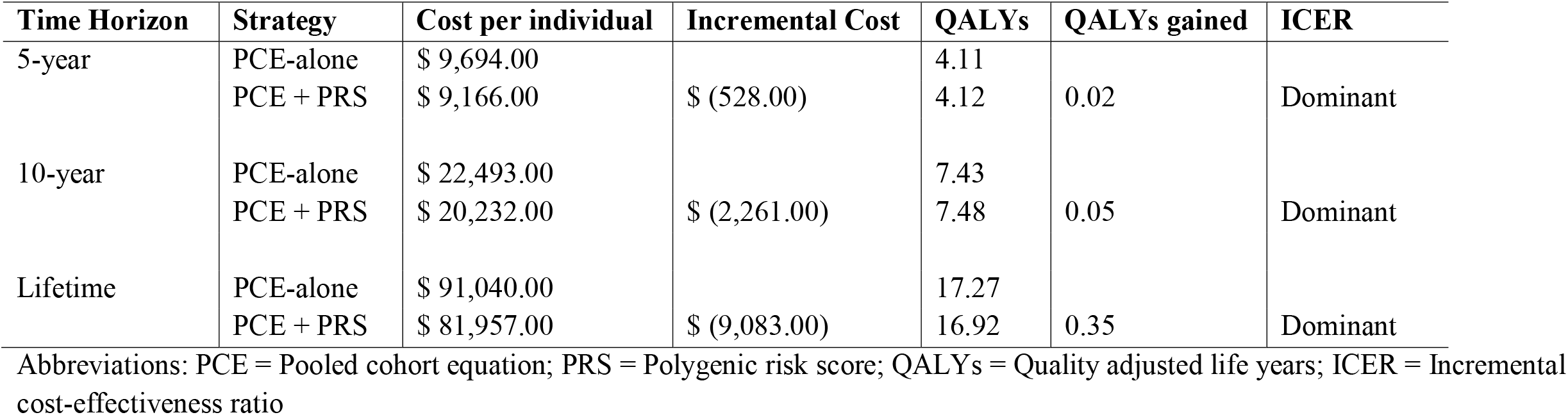
Base case cost-effectiveness results.

**Table 3.** Proportion of the cohort with CAD or Stroke stratified by strategy.

| Time Horizon | PCE |  | PCE + PRS |  | (PCE vs PCE+PRS) |  | CAD or Stroke cases prevented<br>(Per 1000 persons) | Cost per CAD or Stroke case prevented |
| --- | --- | --- | --- | --- | --- | --- | --- | --- |
|  | CAD | Stroke | CAD | Stroke | CAD | Stroke |  |  |
| 5 Years | 10.59 | 3.48 | 8.41 | 2.97 | 2.19 | 0.51 | 27 | \$ 19,555 |
| 10 Years | 20.09 | 6.44 | 16.23 | 5.58 | 3.85 | 0.86 | 47 | \$ 48,106 |
| Lifetime | 57.29 | 16.89 | 50.20 | 15.72 | 7.09 | 1.17 | 83 | \$ 109,433 |

Abbreviations: PCE = Pooled cohort equation; PRS = Polygenic risk score; CAD = coronary artery disease

### One- and two-way sensitivity analysis

Results for the one-way sensitivity in a 5 year time horizon are presented in **Figure 2**. Findings were robust (ICER < $ 50,000 WTP threshold) to variations in parameter inputs with cost of PRS testing having the largest impact on the ICER. PCE+PRS remained cost-saving and dominant in the 10 year and lifetime time horizons regardless of the parameter inputs’ ranges (supplementary material, Figures S1 and S2). In two-way sensitivity, PCE+PRS was more cost-effective at higher statin adherence levels, lower cost of PRS testing and longer analytic time horizons (supplementary material, Figures S7-S9). With perfect (100%) statin adherence, PCE+PRS was cost-effective when the cost of PRS testing was less than $550, $1300, and $6500 in the 5 year, 10 year, and lifetime time horizons respectively.

**Figure 2:**
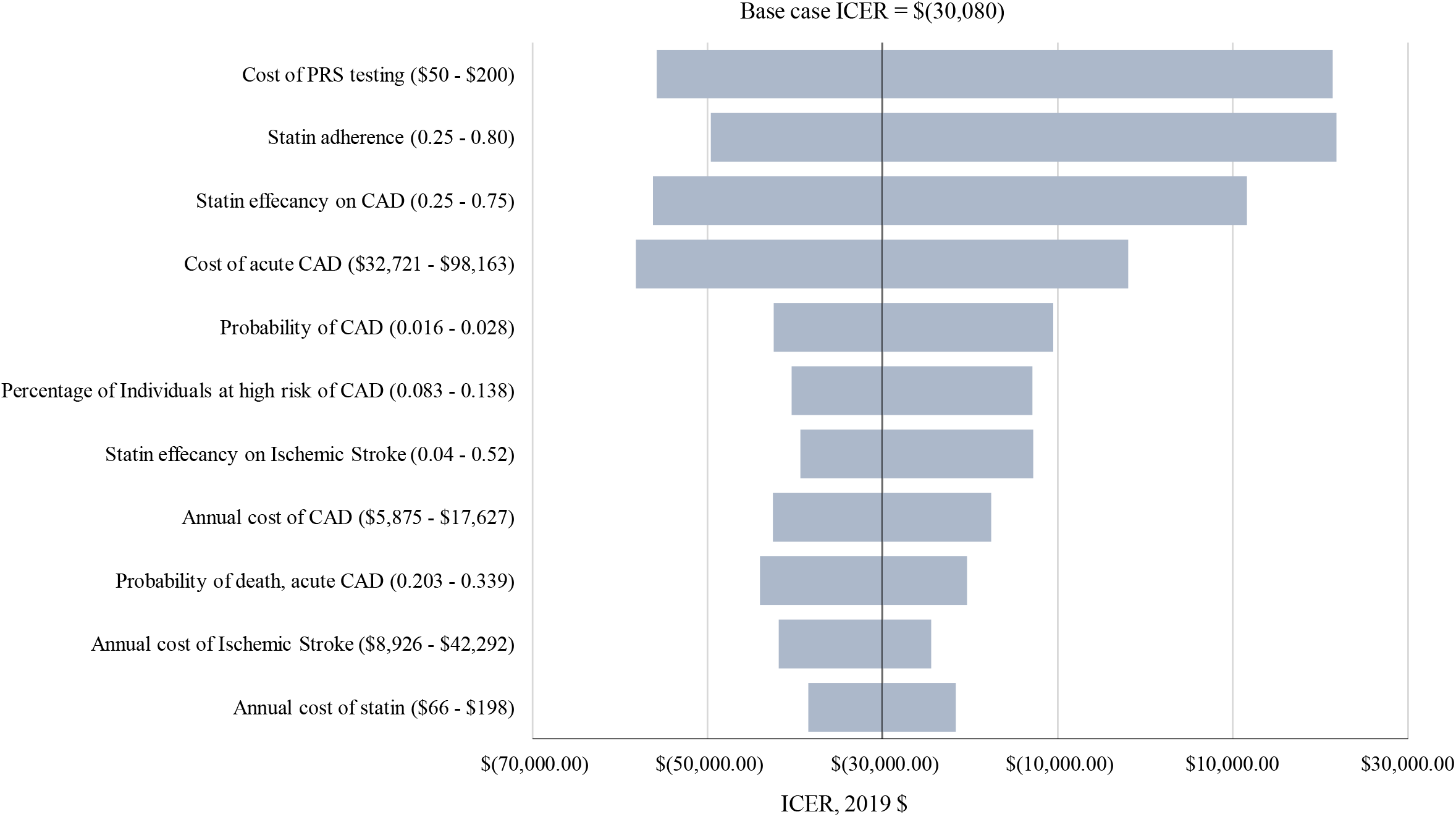
Key variables impacting the ICER for PCE-alone vs PCE+PRS considering a 5-year time horizon.

### Probabilistic sensitivity analysis

We performed 10,000 Monte Carlo simulations and nearly all the ICER points in the cost-effectiveness planes (**Figure 3-Panel A**) fell below the WTP threshold of $50,000 and in the southeast quadrant, indicating that PCE+PRS was effective, cost-saving, and cost-effective compared to PCE-alone. Further, PCE+PRS was more cost-saving and cost-effective with increased time analytic horizons (supplementary material, Figures S3-S6). The probability of PCE+PRS being cost-effective was nearly one at a WTP threshold of $50,000 in 5 year (**Figure 3-Panel B**), 10 year, and lifetime time horizons (supplementary material, Figures S5 and S6).

**Figure 3:**
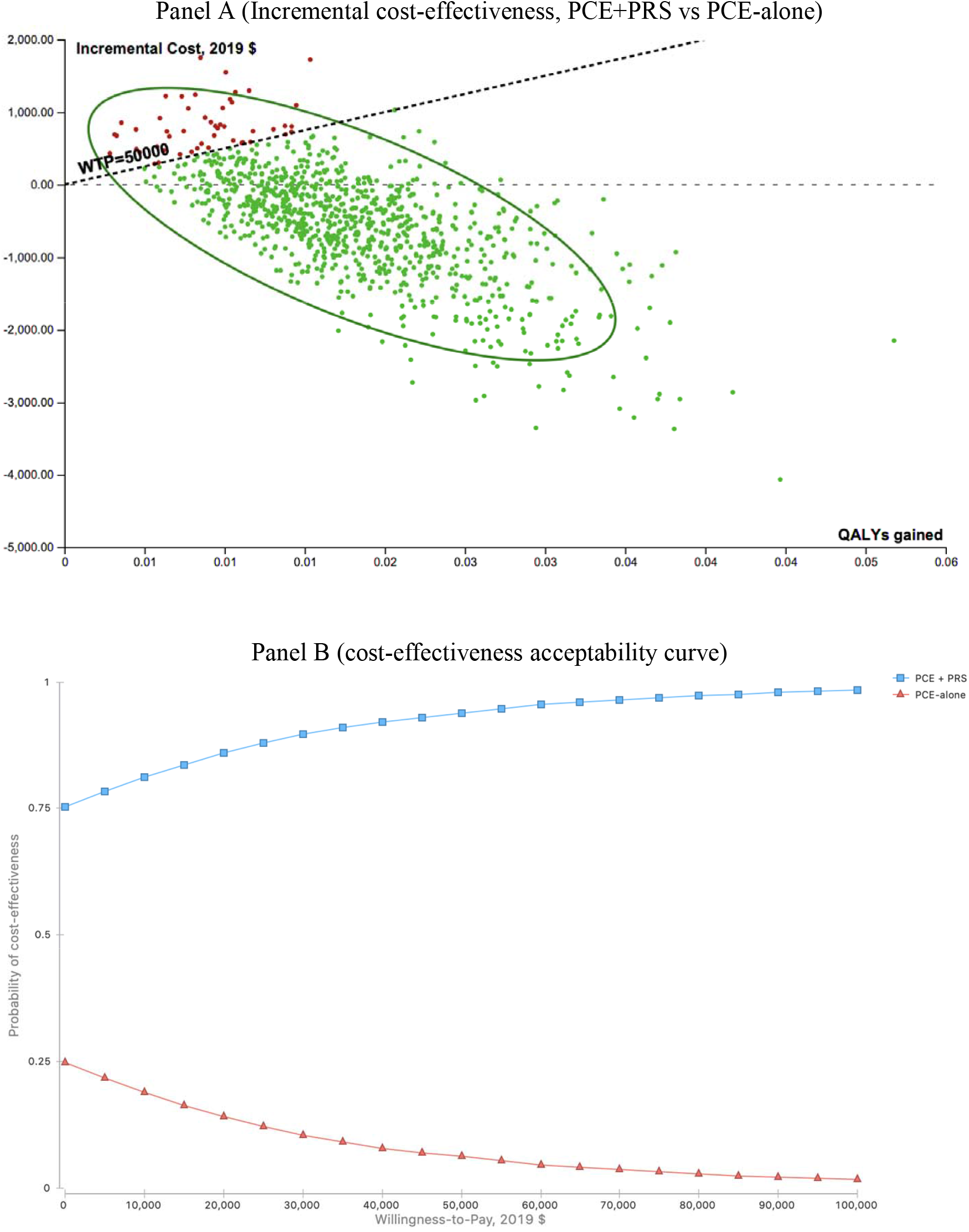
probabilistic sensitivity analysis results for a 5 year time horizon.

### Scenario analysis

When we included the cost of an additional primary care visit, PCE+PRS was cost-effective (ICER = $71,800) compared to PCE-alone at $100,000 WTP threshold in the 5 year time horizon. However, PCE+PRS was dominant compared to PCE-alone in the 10 year and lifetime time horizons (supplementary material, table S2). PCE+PRS was more cost-saving and effective (supplementary material, table S3) than in the base case after adjusting for age in the risk of CAD. In the best-case scenario with perfect adherence to statin therapy and age-adjusted risk of CAD, the PCE+PRS strategy was more cost-saving, cost-effective and prevented more acute CAD and ischemic stroke events compared to the base-case (supplementary material, table S4 and S5). Variations in the cohort start age indicated that implementing PRS at a younger age would save the health system more money, especially in the long run (**Figure 4**).

**Figure 4:**
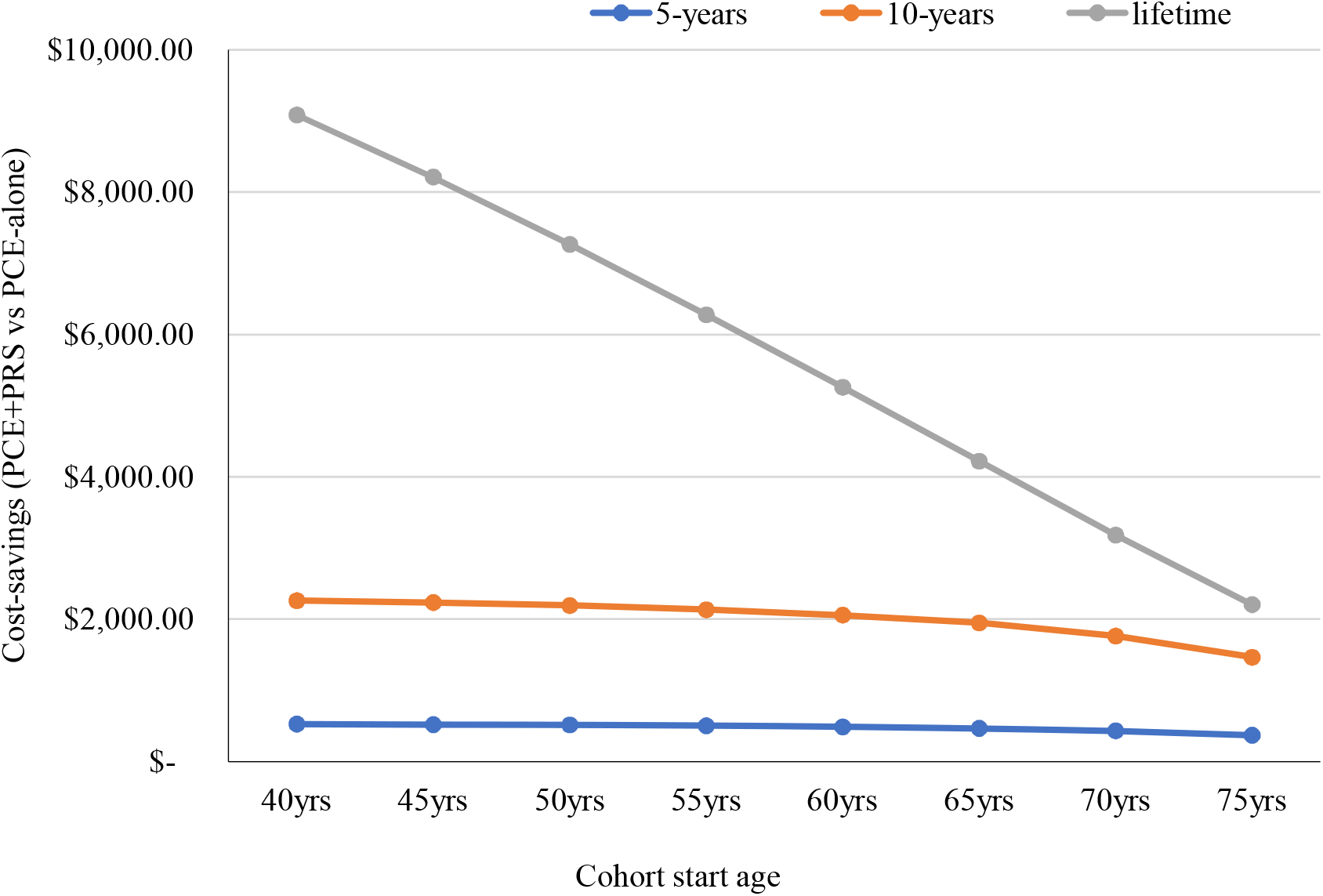
Cost-savings per high-risk individual, PCE+PRS vs PCE-alone.

## Discussion

We developed a Markov model to examine the cost-effectiveness of integrating CAD-PRS into the PCE to identify high-risk individuals who are undetected by current clinical guidelines — PCE-alone. Our results support the growing body of evidence of the health and economic benefits of using PRS to determine an individual’s risk for complex chronic conditions, including cardiovascular diseases, and for informing preventive therapy decisions.^18,19^ Implementing CAD-PRS as an additional risk enhancing factor to reclassify individuals identified with borderline or intermediate risk of ASCVD with the PCE could save more than $500, $2,200, and $9,000 per additional high-risk individual identified in 5 year, 10 year, and lifetime time horizons, respectively, while at the same time improving quality of life.

The cost of PRS testing and statin adherence were the main drivers of the cost-effectiveness of PCE+PRS, particularly in the 5 year time horizon, although the probability of death from acute CAD and the cost of non-fatal acute CAD and ischemic stroke in the 10 year and lifetime time horizons were also important, consistent with previous studies.^18,19^ For the 5 year time horizon, PCE+PRS was cost-saving when the cost of PRS testing and the percentage of the cohort adherence to statin were less than $155 and 35%, respectively, but remained cost-effective at all parameter values. Nevertheless, when viewed across the lifetime of the cohort, PCE+PRS was both cost-saving and cost-effective regardless of parameter estimate uncertainty ranges (supplementary material, Figures S1 and S2).

Our findings highlight both short- and long-term health and economic benefits of integrating CAD-PRS into risk assessments for ASCVD. More than 27, 47 and 83 acute CAD and ischemic stroke events are prevented by the PCE+PRS compared to PCE-alone per 1,000 persons in 5 year, 10 year, and lifetime time horizons, respectively. As a result, health systems could save an average of $19,500 - $109,000 per acute CAD or ischemic stroke event averted between the 5 year and lifetime time horizons, suggesting that the longer the outlook, the more beneficial implementing PRS becomes. Furthermore, from a societal perspective, PCE+PRS may be even more cost-saving when loss of productivity from CAD or stroke events is also considered.^39,40^

CAD-PRS has greater benefits when implemented in young adults given the limitations of traditional risk factors in identifying the risk of ASCVD at a younger age.^6^ Our results indicate higher QALYs gained and cost-savings when PCE+PRS was implemented in a 40-vs 75-year-old cohort, primarily due to the long term benefits from prevention of adverse health outcomes in young adults (Figure 4). Previous work has demonstrated that individuals who receive their genetic results are more likely to report behavior changes.^41–43^ For example, in a Finnish study patients who received information on their genetic risk for cardiovascular disease were more likely to stop smoking, reduce their weight and visit their doctor more.^42^ Therefore, implementing CAD-PRS in young adults has the corollary of improving quality of life in the long run.

Whilst our findings are broadly comparable with previous work, they also provide important novel insights on the efficiency of precision medicine, particularly in primary prevention for CAD. In one study, genetic testing was more beneficial when targeting individuals where traditional risk factors do not provide an accurate risk assessment.^18^ Combining traditional risk factors with genetic testing for a segment of individuals with 17-22% 10-year risk reduced the average cost of treating cardiovascular diseases per individual in a population of 100,000 adults by 2.54 euros in a 10-year follow-up compared to using traditional risk factors on their own.^18^ In our study, PCE+PRS saved more than $2,300 in expected costs with an extra 0.05 QALYs gained per additional high-risk individual identified, compared to PCE-alone after 10 years. This translated to an average of $90 saved per 40-year-old individual in the primary prevention population. Our study identified more high risk individuals since genetic testing was performed on a larger percentage (36%)—those with 5% to <20% 10-year risk—of the primary prevention population compared to the optimization approach used by Hynninen et al. (2019) where genetic testing was performed on only 3% of the population—those with 17% to <22% 10-year risk based on traditional risk factors—and excluding those with <10% 10-year risk from any preventive therapy.^18^ In a recent US-based study, genetic testing on individuals with low to borderline (2.5% to 7.5%) 10 year risk was found to not be cost-effective in informing statin therapy decisions.^19^ However, these authors only considered individuals with low and borderline 10 year risk and applied genetic testing to classify high-risk individuals for preventive care, whereas guidelines recommend that additional risk-enhancing factors should be used on individuals at borderline or intermediate 10 year risk (5% to <20% 10-year risk).^7^ Accordingly, in this study we implemented PCE+PRS on individuals with borderline/intermediate risk and demonstrate enhanced health and economic outcomes when focusing on this sub-group.

### Limitations

Our study has several limitations. First, alternative preventive therapies were not considered and individuals that experienced statin therapy side effects were assumed to not be on any preventive therapy. Even though some studies have shown efficacy of alternative preventive therapies (e.g., PCSK9),^44^ statin therapy remains the recommended first line preventive care for cardiovascular diseases in the US.^1^ Second, we assumed high-risk individuals remained unidentified under the PCE-alone strategy throughout the analytic time horizons and did not initiate any preventive therapy, which may not always be the case in a real-world setting since age is a strong risk factor of CAD and may inform prevention decisions, particular over the course of a lifetime. Third, our model examined prevention of the first acute CAD and ischemic stroke events and not subsequent events. Studies have shown that patients with high PRS may get more health benefits from some prevention therapies, thus improving outcomes in secondary prevention.^45^ Fourth, some data were based on retrospective and observational studies due to lack of prospective studies examining the effectiveness of incorporating PRS into heart health checks.^46^ Fifth, due to data limitations, our study cohort combined individuals with borderline and intermediate risk although guidelines recommend moderate-intensity statin therapy for those with borderline risk and an additionally risk enhancing factor and high-intensity statin therapy for those with intermediate risk and additionally risk enhancing factor.^7^ However, our findings are conservative since we assumed moderate-intensity statin therapy. Finally, we assumed that patient behavior did not change over the analytical time horizon, whilst there is some evidence that genetic testing is associated with positive changes in patient behavior.^41–43^

## Conclusion

To inform preventive care decisions in a real-world clinical setting, we developed a Markov model to examine the cost and health outcomes from implementing CAD-PRS among individuals with borderline or intermediate PCE 10-year risk of ASCVD. We found that implementing PRS in the primary prevention of CAD is an efficient use of resources and saves money for health systems.

## Data Availability

Data are available from the literature

