## Supplementary material for "Integrating a Polygenic Risk Score for Coronary Artery Disease as a Risk Enhancing Factor in the Pooled Cohort Equation is Cost-effective in a US Health System"

**Title**

^1^Allelica, Inc

^2^Illumina, Inc

**Overview**

The supplementary material includes tables and figures from the additional analysis and sensitivity analysis

**Table S1: Expected costs breakdown per high-risk individual for the base case analysis cost-effectiveness results**

| **Cost component** | **5-year time horizon** | | | **10-year time horizon** | | | **Lifetime time horizon** | |
| --- | --- | --- | --- | --- | --- | --- | --- | --- |
|  | **PCE-alone** | **PCE + PRS** | **PCE-alone** | | **PCE + PRS** | **PCE-alone** | | **PCE + PRS** |
| PRS testing | $ - | $ 903.34 | $ - | | $ 903.34 | $ - | | $ 903.34 |
| Statin Therapy | $ - | $ 295.11 | $ - | | $ 517.08 | $ - | | $ 1,059.02 |
| Diabetes | $ - | $ 106.66 | $ - | | $ 291.79 | $ - | | $ 1,179.64 |
| Hemorrhagic stroke | $ - | $ 26.32 | $ - | | $ 63.98 | $ - | | $ 230.13 |
| Myopathy | $ - | $ 13.21 | $ - | | $ 35.99 | $ - | | $ 144.85 |
| Ischemic stroke | $ 2,335.98 | $ 1,989.35 | $ 5,808.77 | | $ 4,997.22 | $ 21,342.27 | | $ 19,205.53 |
| Myocardial Infarction | $ 7,358.35 | $ 5,832.44 | $ 16,684.11 | | $ 13,422.32 | $ 69,698.08 | | $ 59,234.01 |
| **Total costs** | **$ 9,694.33** | **$ 9,166.44** | **$ 22,492.88** | | **$ 20,231.71** | **$ 91,040.35** | | **$ 81,956.52** |

Abbreviations: PCE = Pooled cohort equation; PRS = Polygenic risk score

Majority of the costs are attributable to treatment for myocardial infarction and ischemic stroke

**Table S2: Cost-effectiveness results with the cost of an additional primary care visit included in the PCE+PRS strategy**

| Time Horizon | Strategy | Cost per individual | Incremental Cost | QALYs | QALYs gained | ICER |
| --- | --- | --- | --- | --- | --- | --- |
| 5-year | PCE-alone | $ 9,694.00 |  | 4.11 |  |  |
|  | PCE + PRS | $ 10,955.00 | $ 1,261.00 | 4.13 | 0.02 | $ 71,800.00 |
| 10-year | PCE-alone | $ 22,493.00 |  | 7.43 |  |  |
|  | PCE + PRS | $ 22,020.00 | $ (473.00) | 7.48 | 0.05 | $ (8,700.00) |
| Lifetime | PCE-alone | $ 91,040.00 |  | 16.92 |  |  |
|  | PCE + PRS | $ 83,745.00 | $ (7,295.00) | 17.27 | 0.35 | $ (20,700.00) |

Abbreviations: PCE = Pooled cohort equation; PRS = Polygenic risk score; QALYs = Quality adjusted life years; ICER = Incremental cost-effectiveness ratio

Table S2 shows cost-effectiveness results after including the cost an extra primary care visit to explain to the patient the benefits of PRS testing. At a willingness to pay threshold of $50,000, implementing PRS was not cost-effective in a 5-year time horizon but cost-saving and cost-effective in 10-year and lifetime time horizon.

**Table S3: Cost-effectiveness results with age-adjusted risk for CAD**

| **Time Horizon** | **Strategy** | **Total Cost** | **Incremental Cost** | **QALYs** | **QALYs gained** | **ICER** |
| --- | --- | --- | --- | --- | --- | --- |
| 5-year | PCE-alone | $ 10,298.00 |  | 4.10 |  |  |
|  | PCE + PRS | $ 9,651.00 | $ (647.00) | 4.42 | 0.02 | $ (35,000.00) |
| 10-year | PCE-alone | $ 25,223.00 |  | 7.39 |  |  |
|  | PCE + PRS | $ 22,486.00 | $ (2,737.00) | 7.45 | 0.06 | $ (46,000.00) |
| Lifetime | PCE-alone | $ 112,885.00 |  | 16.11 |  |  |
|  | PCE + PRS | $ 103,717.00 | $ (9,168.00) | 16.47 | 0.36 | $ (25,530.00) |

Abbreviations: PCE = Pooled cohort equation; PRS = Polygenic risk score; QALYs = Quality adjusted life years; ICER = Incremental cost-effectiveness ratio; CAD = coronary artery disease

Table S3 shows cost-effectiveness results after adjusting for age in the risk of CAD. PCE+PRS is more cost-saving and cost-effective in all time horizons after adjusting for age in the risk of CAD.

**Table S4: Cost-effectiveness results for best-case scenario**

| Time Horizon | Strategy | Total Cost | Incremental Cost | QALYs | QALYs gained | ICER |
| --- | --- | --- | --- | --- | --- | --- |
| 5-year | PCE-alone | $ 10,298.00 |  | 4.10 |  |  |
|  | PCE + PRS | $ 8,071.00 | $ (2,227.00) | 4.14 | 0.04 | $ (60,000.00) |
| 10-year | PCE-alone | $ 25,223.00 |  | 7.39 |  |  |
|  | PCE + PRS | $ 18,679.00 | $ (6,545.00) | 7.52 | 0.12 | $ (54,000.00) |
| Lifetime | PCE-alone | $ 112,885.00 |  | 16.11 |  |  |
|  | PCE + PRS | $ 91,171.00 | $ (21,714.00) | 16.92 | 0.81 | $ (27,000.00) |

Abbreviations: PCE = Pooled cohort equation; PRS = Polygenic risk score; QALYs = Quality adjusted life years; ICER = Incremental cost-effectiveness ratio; CAD = coronary artery disease

Table S4 shows cost-effectiveness results for the best-case scenario (age-adjusted risk of CAD and 100% adherence to statin therapy). PCE+PRS can save the health system more than $2,000; $6,500; and $21,000 per additional high-risk individual identified when conditions of the best-case scenario are full filled.

**Table S5. Proportion of the cohort that get acute CAD or ischemic stroke, stratified by strategy (PCE-alone, PCE+PRS)**

| **Time Horizon** | **PCE-alone** | | **PCE + PRS** | | **(PCE vs PCE+PRS)** | | **CAD or Stroke**  **events prevented**  **(Per 1000 persons)** | **Cost per CAD or Stroke**  **event prevented** |
| --- | --- | --- | --- | --- | --- | --- | --- | --- |
|  | CAD | Stroke | CAD | Stroke | CAD | Stroke |  |  |
| 5 Years | 11.64 | 3.47 | 6.81 | 2.45 | 4.83 | 1.02 | 59 | $ 38,068 |
| 10 Years | 24.48 | 6.32 | 15.04 | 4.62 | 9.44 | 1.70 | 111 | $ 58,752 |
| Lifetime | 84.07 | 12.70 | 73.69 | 11.32 | 10.38 | 1.38 | 118 | $ 184,642 |

Abbreviations: PCE = Pooled cohort equation; PRS = Polygenic risk score; CAD = coronary artery disease

Table S5 shows events of CAD or stroke by strategy. More events of CAD or stroke are prevented in the best-case scenario which saves the health system more than $38,000; $58,000; and $184,000 per event prevented in 5 years, 10 years and lifetime time horizon.

**Table S6: Total costs per high-risk individual**

| **Cohort start age** | **5-year time horizon** | | **10-year time horizon** | | **Lifetime time horizon** | |
| --- | --- | --- | --- | --- | --- | --- |
|  | **PCE-alone** | **PCE+PRS** | **PCE-alone** | **PCE+PRS** | **PCE-alone** | **PCE+PRS** |
| 40yrs | $ 9,694.00 | $ 9,166.00 | $ 22,493.00 | $ 20,232.00 | $ 91,040.00 | $ 81,957.00 |
| 45yrs | $ 9,662.00 | $ 9,142.00 | $ 22,307.09 | $ 20,071.63 | $ 80,970.88 | $ 72,759.79 |
| 50yrs | $ 9,610.17 | $ 9,094.55 | $ 21,995.49 | $ 19,803.24 | $ 70,654.10 | $63,388.58 |
| 55yrs | $ 9,528.12 | $ 9,024.47 | $ 21,570.20 | $ 19,436.98 | $ 60,401.94 | $ 54,126.05 |
| 60yrs | $ 9,423.67 | $ 8,935.28 | $ 21,030.14 | $ 18,971.82 | $ 50,398.40 | $ 45,139.90 |
| 65yrs | $ 9,284.57 | $ 8,816.47 | $ 20,238.47 | $ 18,289.65 | $ 40,732.96 | $ 36,512.88 |
| 70yrs | $ 9,042.52 | $ 8,609.73 | $ 18,899.39 | $ 17,135.69 | $ 31,590.84 | $ 28,408.60 |
| 75yrs | $ 8,614.45 | $ 8,244.11 | $ 16,742.03 | $ 15,276.48 | $ 23,416.20 | $ 21,210.83 |

Abbreviations: PCE = Pooled cohort equation; PRS = Polygenic risk score

Table S6 shows total costs per high-risk individual identified by PCE+PRS. PCE+PRS is less costly compared to PCE-alone.

**Table S7: Cost-savings per high-risk individual, PCE+PRS vs PCE-alone**

| **Cohort start age** | **5-year time horizon** | **10-year time horizon** | **Lifetime time horizon** |
| --- | --- | --- | --- |
| 40yrs | $ 528.00 | $ 2,261.00 | $ 9,083.00 |
| 45yrs | $ 520.00 | $ 2,235.46 | $ 8,211.09 |
| 50yrs | $ 515.62 | $ 2,192.26 | $ 7,265.52 |
| 55yrs | $ 503.64 | $ 2,133.22 | $ 6,275.89 |
| 60yrs | $ 488.39 | $ 2,058.31 | $ 5,258.49 |
| 65yrs | $ 468.09 | $ 1,948.82 | $ 4,220.08 |
| 70yrs | $ 432.79 | $ 1,763.70 | $ 3,182.24 |
| 75yrs | $ 370.33 | $ 1,465.54 | $ 2,205.37 |

Abbreviations: PCE = Pooled cohort equation; PRS = Polygenic risk score

Table S7 shows cost-savings per high-risk individual identified by PCE+PRS. PCE+PRS. The health system saves more money when PRS is implemented at younger age as compared to later years. PRS saves more money in the future since the benefits are realized over an individual’s life span.

**Table S8: QALYs per high-risk individual**

| Cohort start age | 5-year time horizon | | 10-year time horizon | | Lifetime time horizon | |
| --- | --- | --- | --- | --- | --- | --- |
|  | PCE-alone | PCE+PRS | PCE-alone | PCE+PRS | PCE-alone | PCE+PRS |
| 40yrs | 4.11 | 4.13 | 7.43 | 7.48 | 16.92 | 17.27 |
| 45yrs | 4.04 | 4.05 | 7.26 | 7.32 | 15.62 | 15.95 |
| 50yrs | 3.95 | 3.97 | 7.06 | 7.12 | 14.24 | 14.53 |
| 55yrs | 3.85 | 3.86 | 6.83 | 6.88 | 12.79 | 13.05 |
| 60yrs | 3.73 | 3.75 | 6.56 | 6.61 | 11.31 | 11.53 |
| 65yrs | 3.61 | 3.62 | 6.24 | 6.29 | 9.78 | 9.96 |
| 70yrs | 3.45 | 3.46 | 5.81 | 5.86 | 8.20 | 8.34 |
| 75yrs | 3.24 | 3.26 | 5.22 | 5.26 | 6.62 | 6.73 |

Abbreviations: PCE = Pooled cohort equation; PRS = Polygenic risk score; QALYs = Quality-adjusted life years

Table S8 shows QALYs per strategy based on the start of the cohort. PCE+PRS has higher QALYs especially when implemented among younger individuals.

**Table S9: QALYs gained per high-risk individual, PCE-alone vs PCE+PRS**

| Cohort start age | 5-year time horizon | 10-year time horizon | Lifetime time horizon |
| --- | --- | --- | --- |
| 40yrs | 0.018 | 0.050 | 0.350 |
| 45yrs | 0.017 | 0.053 | 0.325 |
| 50yrs | 0.017 | 0.052 | 0.294 |
| 55yrs | 0.017 | 0.051 | 0.259 |
| 60yrs | 0.016 | 0.050 | 0.223 |
| 65yrs | 0.016 | 0.049 | 0.184 |
| 70yrs | 0.016 | 0.047 | 0.146 |
| 75yrs | 0.015 | 0.045 | 0.109 |

Abbreviations: PCE = Pooled cohort equation; PRS = Polygenic risk score; QALYs = Quality-adjusted life years

Table S9 shows the QALYs gained by implementing PCE+PRS compared to PCE-alone. More QALYs are gained when PRS is implemented at younger age.

**Figure S1: One-way sensitivity analysis in a 10-year time horizon**

Base case ICER = $(42,034)

ICER, 2019 $

|  | Low | High |
| --- | --- | --- |
| Annual cost of statin ($66 - $198) | -46839.7 | -37227.5 |
| Probability of CAD (0.016 - 0.028) | -45934.8 | -35595.3 |
| Statin effecancy on Ischemic Stroke (0.040 - 0.520) | -47904.7 | -31069.4 |
| Annual cost of Ischemic Stroke ($8,926 - $42,292) | -53664.5 | -36477.8 |
| Statin adherence (0.25 - 0.8) | -48543.7 | -24893.2 |
| Cost of PRS testing ($50 - $200) | -50429.9 | -25241.1 |
| Statin effecancy on CAD (0.25 - 0.75) | -52013.1 | -26369.7 |
| Probability of death, acute CAD (0.203 - 0.339) | -57612.3 | -31447.7 |
| Annual cost of CAD ($5,875 - $17,627) | -55565.2 | -28497.5 |
| Cost of acute CAD ($32,721 - $98,163) | -57240.9 | -26826.4 |

Figure S1 shows that PCE+PRS dominant (ICER< $0) compared to PCE-alone even at parameter limits

**Figure S2: One-way sensitivity analysis in a lifetime time horizon**

Base case ICER = $ (25,788)

ICER, 2019 $

|  | Low value | High value |
| --- | --- | --- |
| Statin adherence (0.25 - 0.8) | -26800.8 | -23074.7 |
| Cost of PRS testing ($50 - $200) | -27070 | -23223.3 |
| Probability of death from acute Ischemic Stroke (0.1 - 0.3) | -27931.3 | -23806.4 |
| Statin effecancy on CAD (0.25 - 0.75) | -27561.2 | -22959.2 |
| Cost of acute CAD ($32,721 - $98,163) | -29427.4 | -22148.1 |
| Annual discount rate (0 - 0.05) | -28599 | -20871.8 |
| Statin effecancy on Ischemic Stroke (0.04 - 0.52) | -28651.5 | -20099.4 |
| Annual cost of Ischemic Stroke ($8,926 - $42,292) | -31593 | -23014.7 |
| Probability of death from acute CAD (0.203 - 0.339) | -33115.9 | -20317.4 |
| Annual cost of CAD ($5,875 - $17,627) | -36622 | -14949.8 |

Figure S2 shows that PCE+PRS dominant (ICER< $0) compared to PCE-alone even at parameter limits

**Figure S3: Incremental cost-effectiveness, PCE+PRS vs PCE-alone in a 10-year time horizon**

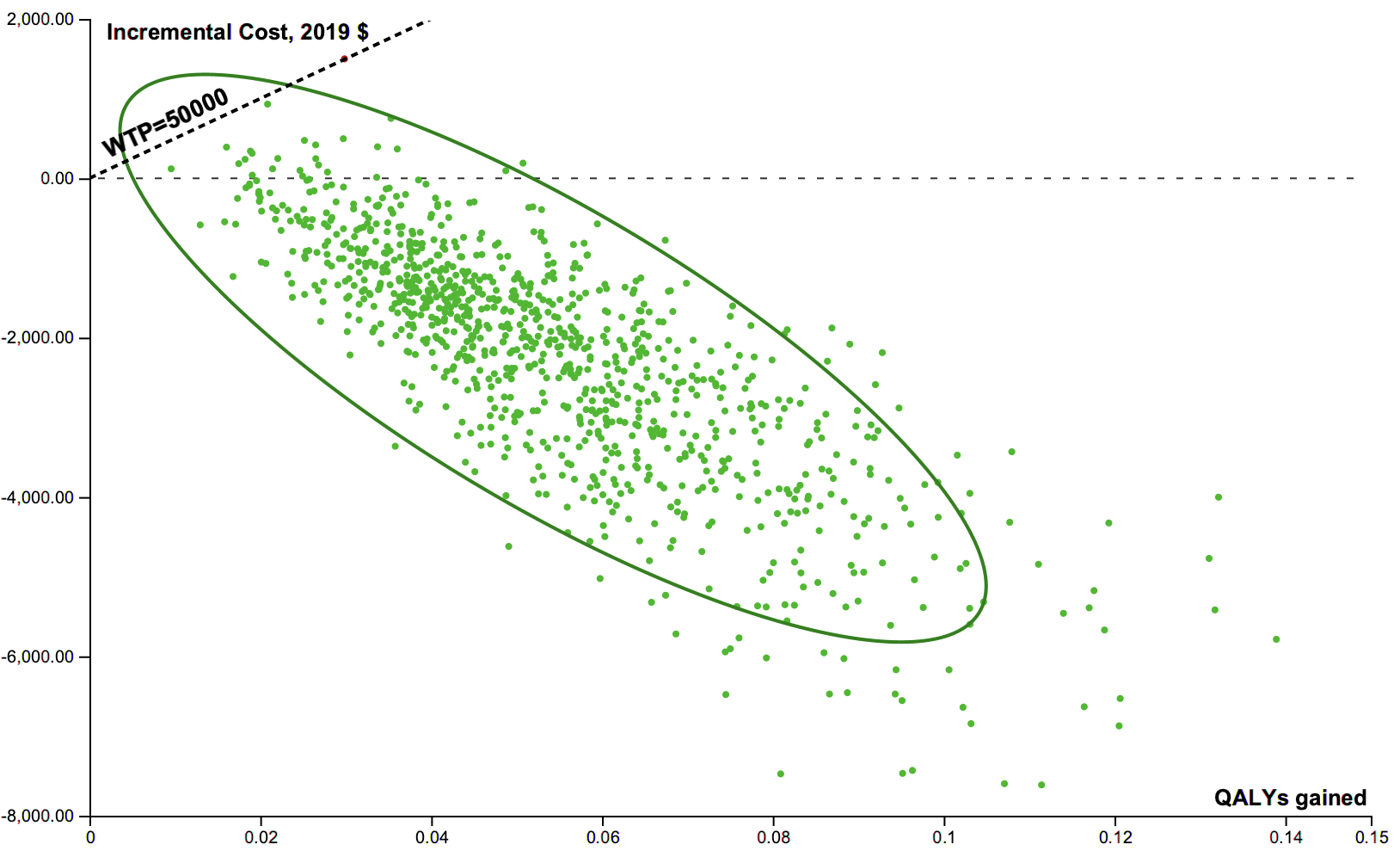

Figure S3 shows a significant majority of the ICER data points from 10,000 simulations fall in the southeast quadrant of the cost-effectiveness plane, indicating that PCE+PRS is always cost-saving and effective compared to PCE-alone.

**Figure S4: Incremental cost-effectiveness, PCE+PRS vs PCE-alone in a lifetime time horizon**

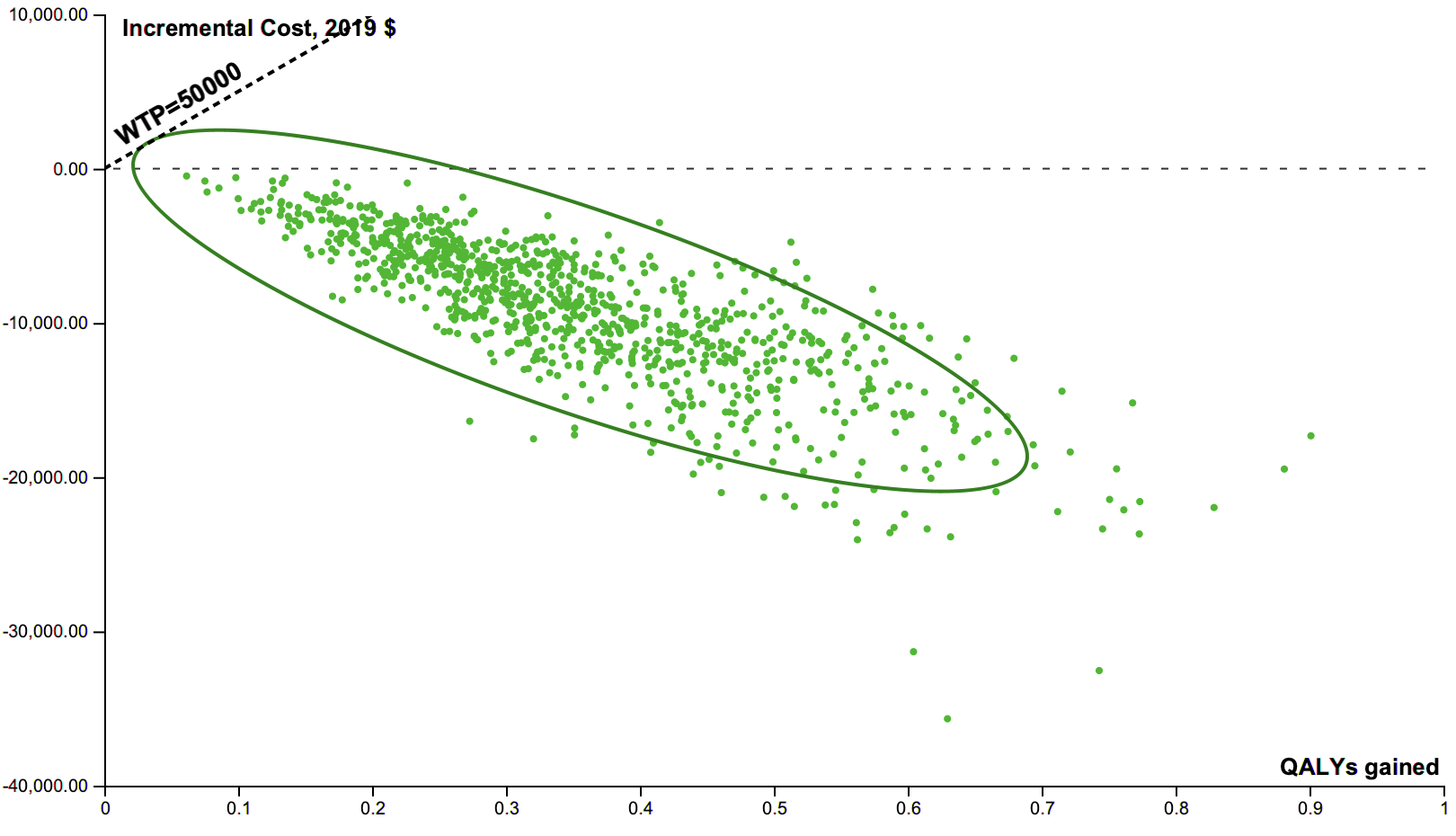

Figure S4 shows all that ICER data points from 10,000 simulations fall in the southeast quadrant of the cost-effectiveness plane, indicating that PCE+PRS is always cost-saving and effective compared to PCE-alone.

**Figure S5: Cost-effectiveness acceptability curve for a 10-year time horizon**

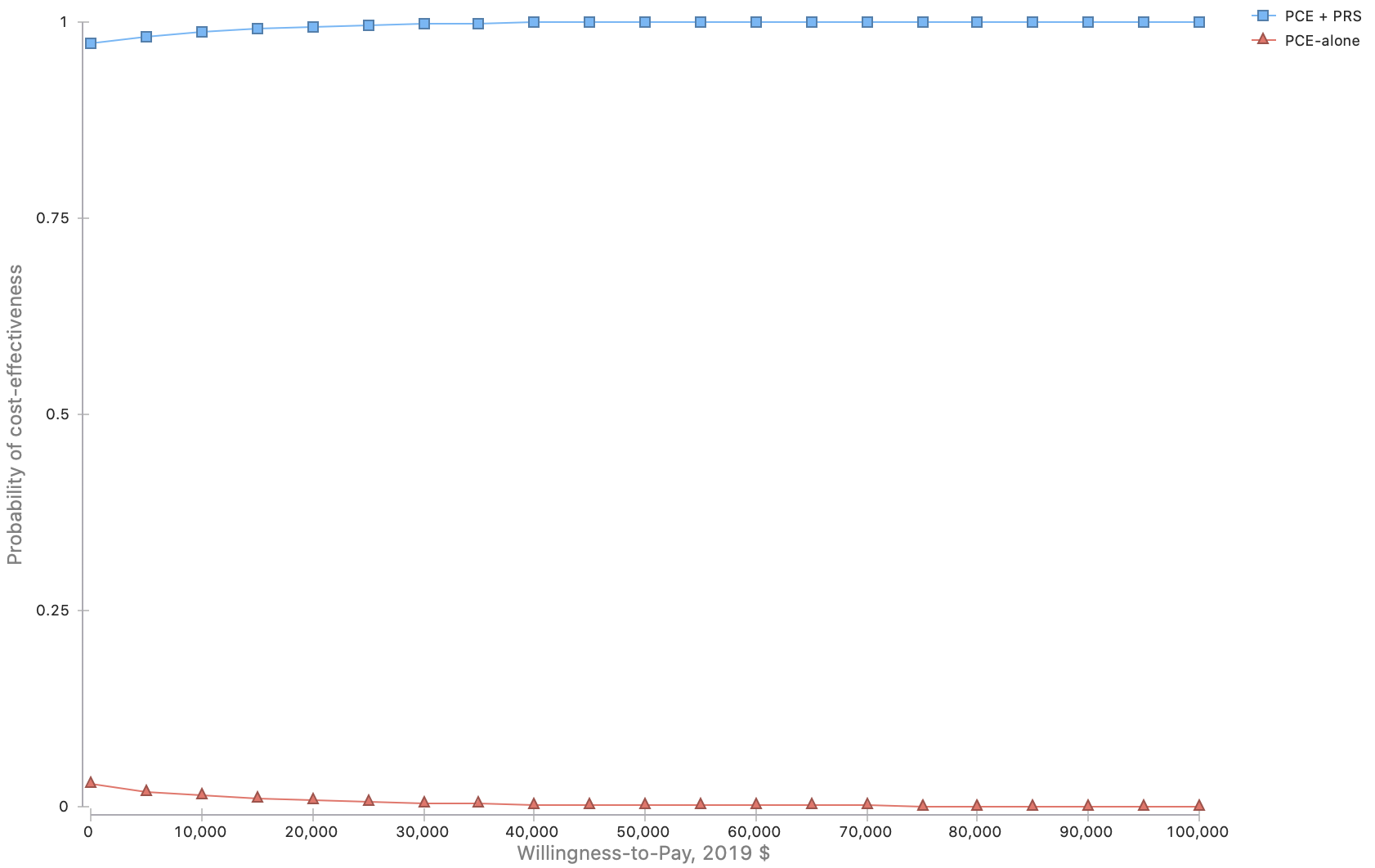

Figure S5 shows that PCE+PRS has a probability of cost-effectiveness of almost 1 compared to PCE-alone at all willingness to pay thresholds.

**Figure S6: Cost-effectiveness acceptability curve for a lifetime time horizon**

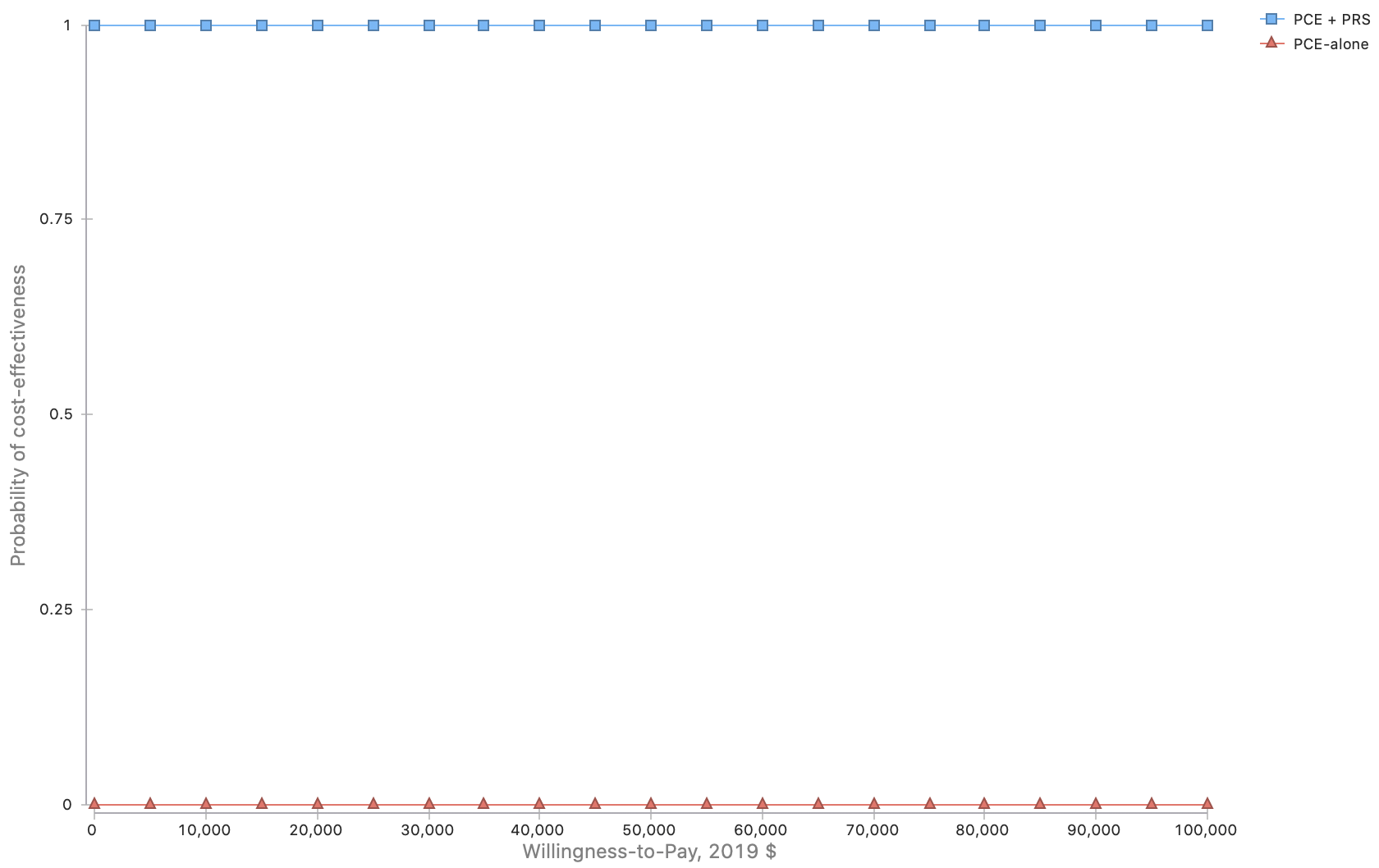

Figure S6 shows that PCE+PRS has a probability of cost-effectiveness of almost 1 compared to PCE-alone at all willingness to pay thresholds.

**Two-way sensitivity analysis**

Figure S7, S8 and S9 show cost thresholds for PRS testing at which implementing PCE+PRS would be a good value for money compared to the current standard clinical practice (PCE-alone) at different statin adherence levels and willingness to pay of $50,000.

**Figure S7: Two-way sensitivity analysis of the cost of PRS testing and statin adherence in a 5-year time horizon**

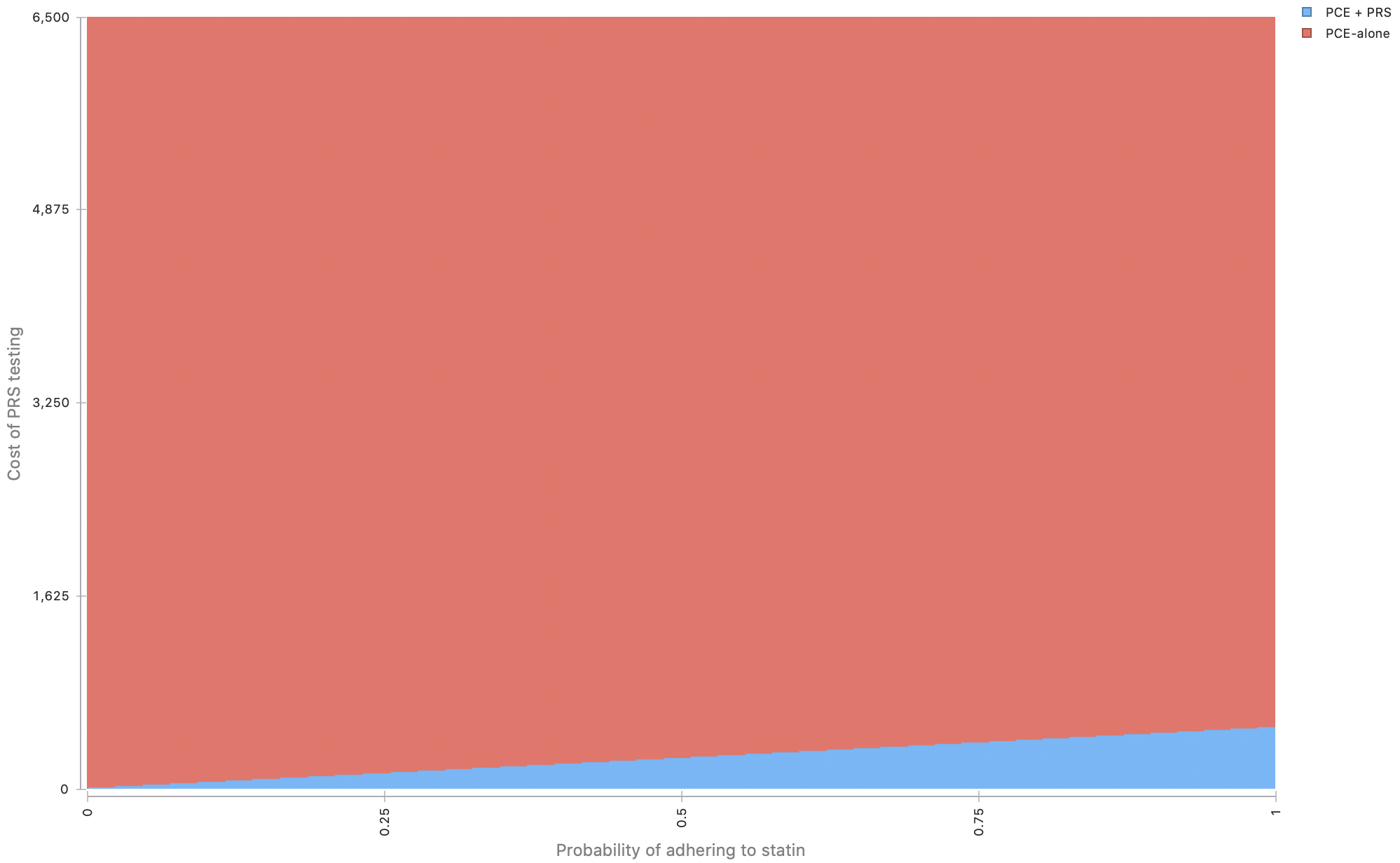

PCE+PRS would be cost-effective compared to PCE-alone at $50,000 willingness to pay when the cost of PRS testing < $5500.

**Figure S8: Two-way sensitivity analysis of the cost of PRS testing and statin adherence in a 10-year time horizon**

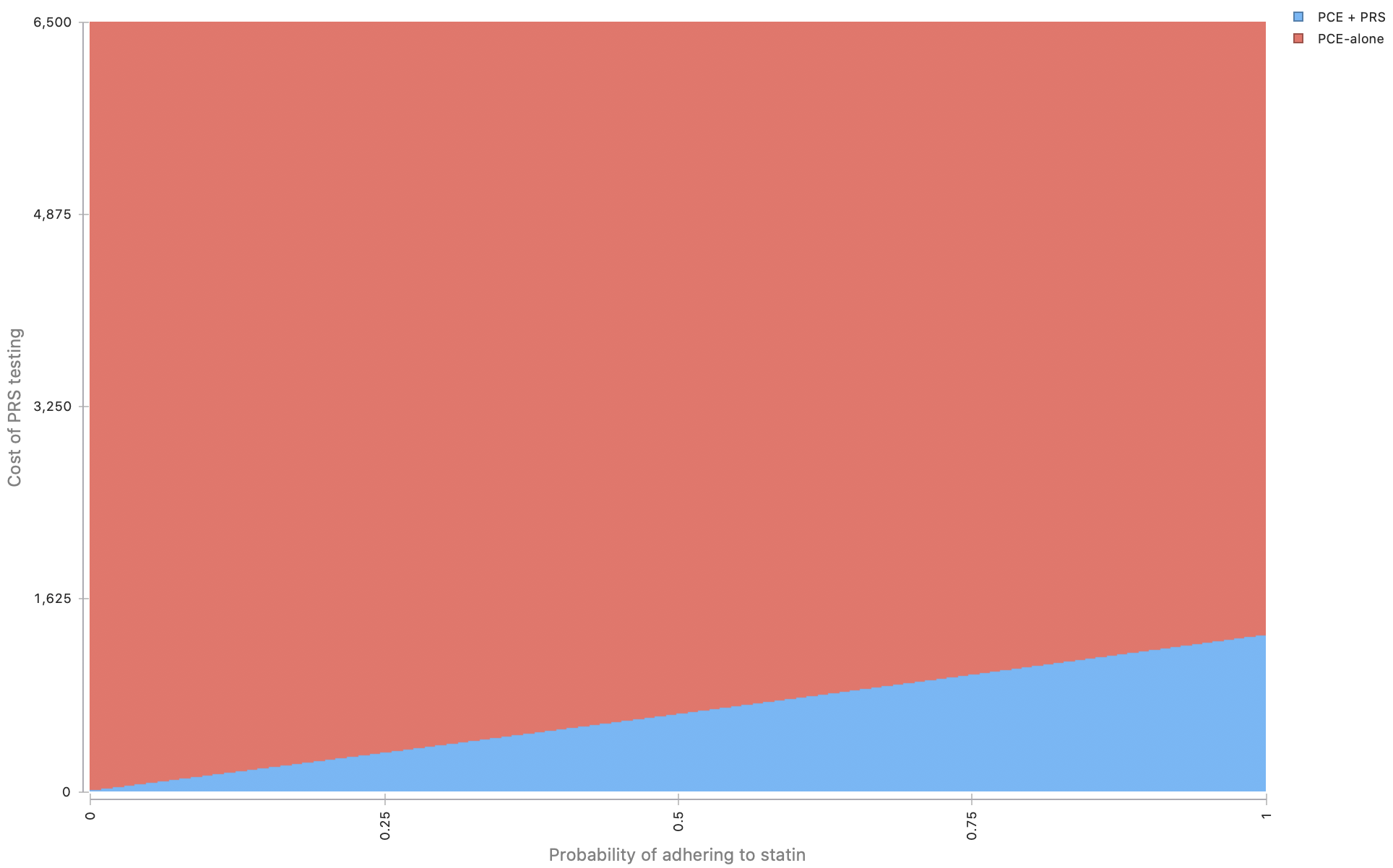

Figure S8 shows that PCE+PRS would be cost-effective compared to PCE-alone at $50,000 willingness to pay when the cost of PRS testing < $1300.

**Figure S9: Two-way sensitivity analysis of the cost of PRS testing and statin adherence in a lifetime time horizon**

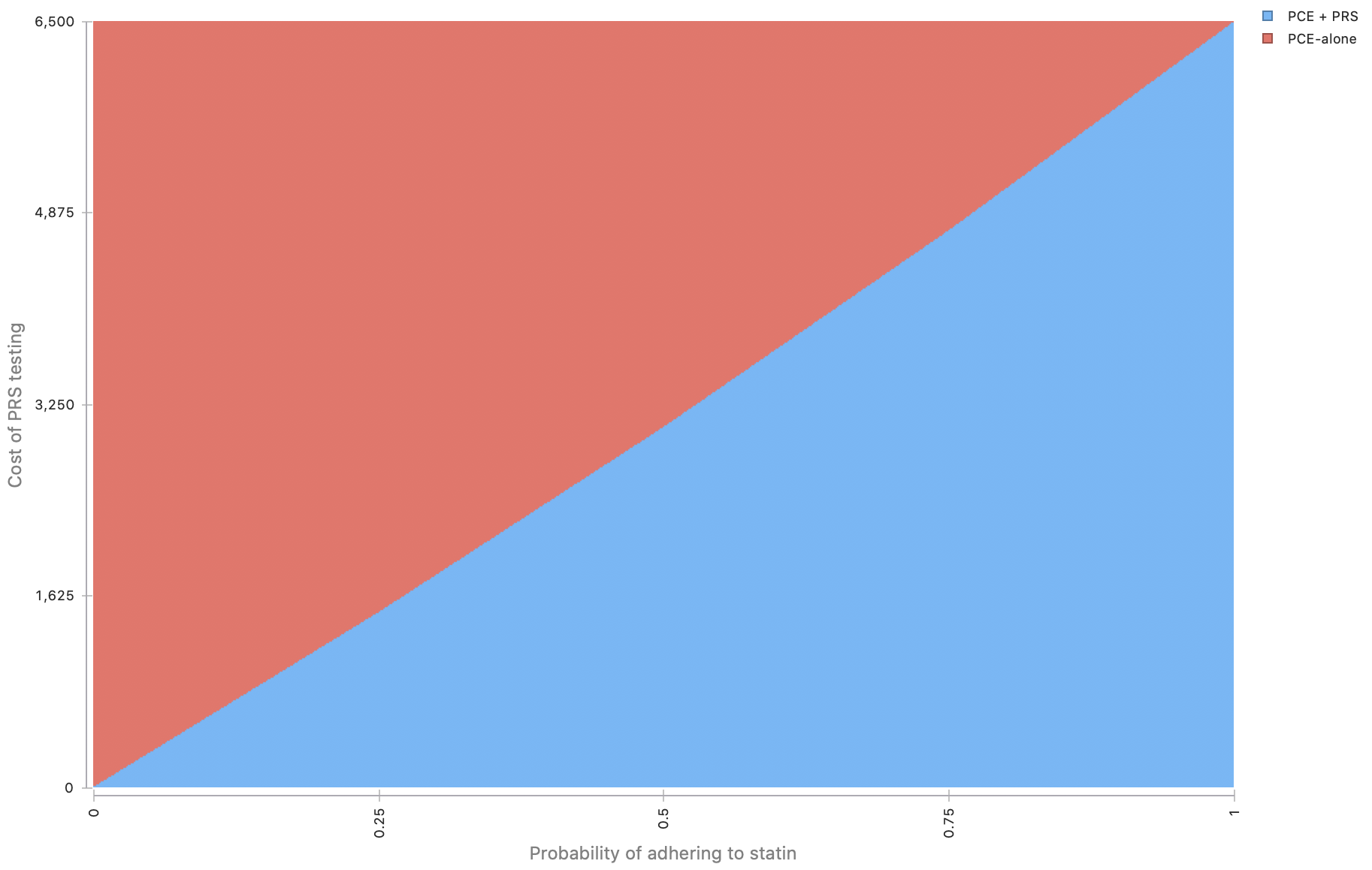

Figure S9 shows that PCE+PRS would be cost-effective compared to PCE-alone at $50,000 willingness to pay when the cost of PRS testing < $6,500.
